# Tinnitus Subtyping with Subgrouping Within Group Iterative Multiple Model Estimation: An Ecological Momentary Assessment Study

**DOI:** 10.1101/2022.02.22.22271338

**Authors:** Jorge Simoes, Jan Bulla, Patrick Neff, Rüdiger Pryss, Steven C. Marcrum, Berthold Langguth, Winfried Schlee

## Abstract

**Background:** Tinnitus is a heterogeneous condition associated with moderate to severe disability, but the reasons why only a subset of individuals is burdened by the condition are not fully clear. Ecological momentary assessment (EMA) allows a better understanding of tinnitus by allowing individualized models and by capturing the fluctuations of tinnitus symptoms and other behavioral dynamics as they occur, and therefore minimizing the risk of recollection bias. The TrackYourTinnitus (TYT) mobile app provides a platform for collecting ecologically valid time series data from tinnitus users and can be used to address questions like how mood, concentration, tinnitus distress, or loudness relate over time. Whether any of those variables have an influence over the next day, that is, whether any of these variables are auto- or cross-correlated, is still unanswered.

**Objectives:** Assess whether behavioral and symptom-related data from tinnitus users from the TYT app auto- and cross-correlate in different time lags, both within and between individuals.

**Methods:** Anonymized data was collected from 278 users of the iOS or Android TYT apps between 2014 and 2020. Tinnitus-related distress, tinnitus loudness, concentration level, overall mood, emotional arousal, and overall stress level were assessed using a 10-point visual analog scale via a daily survey. Auto- and cross-correlations were calculated for participants who used the app for at least 10 consecutive days. Lagged cross-correlation was used to investigate the dynamics of each of these variables over time at the group level, followed by linear regression with elastic net regularization for each user. Additionally, subgrouping within group iterative multiple model estimation (S-GIMME) was used to model the behavioral dynamics at the group, subgroup, and individual levels with data collected from 32 users.

**Results:** No autocorrelation or cross-correlation was observed at the group level between the variables assessed. However, application of the regression models with elastic net regularization identified individualized predictors of tinnitus loudness and distress for most participants, with the models including contemporaneous and lagged information from the previous day. The finding that a subset of users experienced lagged and contemporaneous dynamics was corroborated by the models from S-GIMME. The models had adequate fits, with both contemporaneous and lagged coefficients obtained for most individuals. Two subgroups were identified, the first consisting of users where both contemporaneous and lagged effects were observed, and a second subgroup consisting of users whose dynamics were mainly of contemporaneous effects.

**Discussion:** We showed that tinnitus loudness and tinnitus distress are affected by the contemporaneous and lagged dynamics of behavioral and emotional processes measured through EMA. These effects were seen at the group, subgroup, and individual levels. The relevance EMA and the implications of the insights derived from it for tinnitus care are discussed, especially considering current trends towards the individualization of tinnitus care.

## Introduction

Tinnitus is a condition in which phantom sounds are perceived without a corresponding external stimulus. Those sounds usually take the form of ringing, hissing, or buzzing, but other less common types of perception have also been reported^1,2^. The underlying causes of tinnitus are not fully clear, but auditory pathway deafferentation is commonly recognized as key factor in the etiology of tinnitus^3^. Although tinnitus is generally a benign condition, its bothersome manifestation, which is estimated to affect 1% of the population^4^, can be both debilitating^5^ and costly^6^. Tinnitus can be subdivided into two categories: acute and chronic. The former category describes the rather common phenomenon of phantom sounds being perceived for several seconds or minutes after insult to the auditory system (e.g., listening to loud music); whereas the latter category refers to uninterrupted perception of the phantom sound for at least three months. In its chronic presentation, tinnitus rarely resolves entirely^7,8^.

Available treatments are not effective at suppressing the chronic phantom perception for most sufferers. As a result, most treatment strategies seek to reduce tinnitus-related distress^7^. There is a growing consensus that tinnitus is a heterogeneous condition and that this characteristic may significantly impact both how the condition is experienced by patients and the efficacy of treatments^9,10^. Factors demonstrated to affect treatment outcomes include sociodemographics, personality, and tinnitus characteristics, though many others are currently being investigated^11^. For example, the heritability of tinnitus differs depending on its laterality (i.e., whether sounds are perceived in one or both ears) and the patient’s gender^12^. Furthermore, evidence suggests that personality traits may explain the response to online cognitive behavior therapy treatment^13^, but not acoustic stimulation^14^. It is unclear which factors are related to tinnitus-related disability, as the psychoacoustic properties of tinnitus (e.g., laterality, loudness, pitch, type of perceived sound) do not fully explain the tinnitus distress^9,15^. Therefore, it is of great clinical interest to understand which factors are associated with distress, especially at the individual level.

The advent of minimally intrusive longitudinal sampling methods has allowed researchers and clinicians to develop predictive models at the individual level, while at the same time maximizing the ecological validity of assessments^16^. Not surprisingly, there is growing interest in using ecological momentary assessment (EMA) in the fields of psychology and psychopathology in general^16^ and in tinnitus research in particular. For example, Probst et al.^17^, modeled patterns of daily fluctuation of tinnitus characteristics and identified a mediatorial role of tinnitus loudness on stress, while Pryss et al. used EMA to establish that patients often have recollection bias regarding tinnitus fluctuations throughout the day^18^. However, most of the studies in the tinnitus field using EMA have focused on group-level analysis. A recurring clinical topic is how uniquely tinnitus is experienced by patients, with different triggers leading to higher tinnitus distress and/or loudness in certain patients, but not others. Identification of factors that could lead to personalized interventions is of great clinical utility.

Therefore, the objective of this study was to apply EMA methods to identify factors (see Table 1) that could be associated with tinnitus loudness and distress at both the individual and group levels. Furthermore, EMA lagged data were used to identify whether such factors influence loudness and/or distress on a subsequent day. To achieve these objectives, we first investigated whether states such as tinnitus loudness, distress, and mood impact autocorrelates or cross-correlates throughout subsequent days at the group level. Second, we modeled data at the individual level using linear regressions with elastic net regularization for each unique time series with tinnitus distress and tinnitus loudness as dependent variables. Third, we used subgrouping within group iterative multiple model estimation (S-GIMME) to obtain unique models for each participant on contemporaneous and lagged effects between the variables collected with EMA.

**Table 1.**
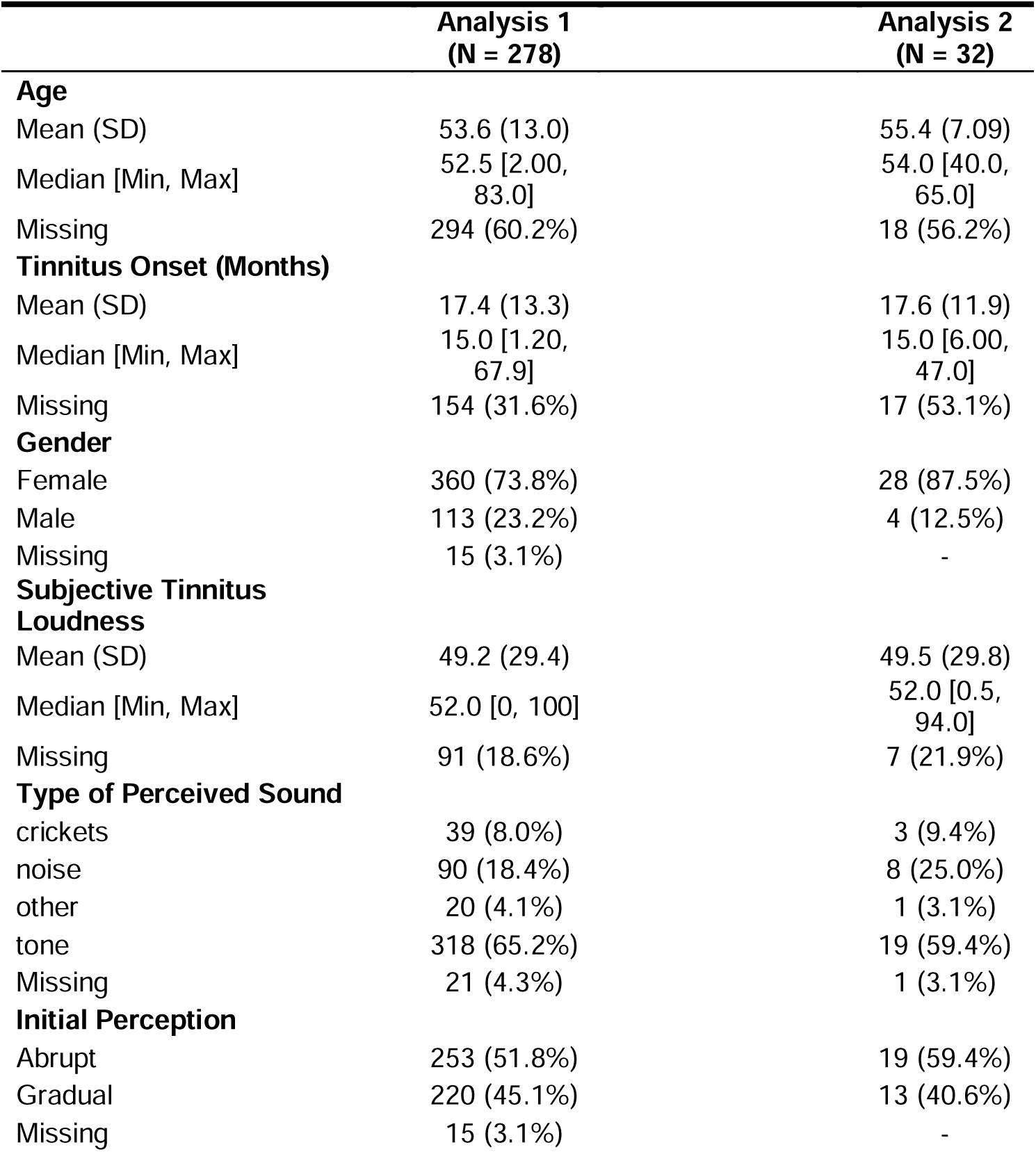
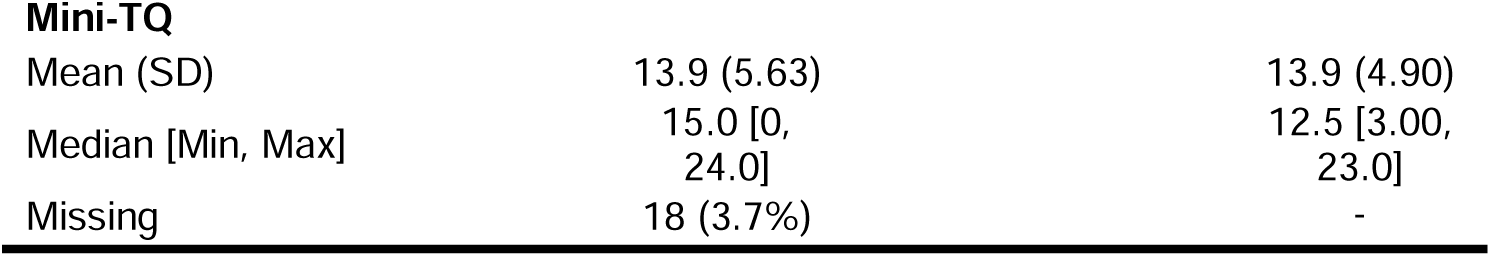
Sample demographics collected during registration in the APP. sample 1 provided sequences with at least 10 days of uninterrupted usage, sample 2 provided sequences with at least 60 days of uninterrupted usage

## Methods

### Data Preparation

The data analyzed in this study were collected between 2014 and 2020 from the ‘TrackYourTinnitus’ (TYT) app, which is freely available on both Android and iOS mobile devices (available as TrackYourTinnitus). After registration, participants complete an 8-question survey with questions related to their current perception of tinnitus and mood. Users are free to use the app for an indeterminate time and without frequency restrictions. An overview of the variables included in this study is available in Table 1. Two variables were excluded from this study: “do you perceive your tinnitus right now?” and “do you feel irritable right now?”, as those questions were binary and continuous variables were necessary to obtain auto-and cross-correlations. The remaining variables were rated on a scale of 0 to 10 using a self-assessment manikin. Although providing daily information about tinnitus could increase user distress by repeatedly directing their attention to their tinnitus, previous work has suggested that using the TYT app does not harm users^19^. Informed consent was obtained from users to have their data anonymously used for scientific purposes. The study was approved by the Ethics Committee of the Faculty of Medicine of the University of Regensburg (Study approval number 15-101-0204).

During registration, users completed two questionnaires: the Mini-Tinnitus Questionnaire^20^ and the Tinnitus Sample Case History Questionnaire, TSCHQ^21^. In addition, users responded to a question about their worst tinnitus-related symptom. Mini-TQ is commonly used in clinical trials and ambulatory assessment as a screening tool for tinnitus-related distress. The questionnaire possesses good psychometric properties (correlation > 0.9 with the original 52-item Tinnitus Questionnaire, test-retest reliability of 0.89, and Cronbach’s alpha of 0.9) and consists of 12 questions. The second questionnaire is part of an international effort to standardize data collection and reporting in tinnitus research and is also a standard screening tool. The TSCHQ consists of 34 questions related to tinnitus characteristics (e.g., the type of perceived sounds, duration of tinnitus, subjective loudness), life history (e.g., whether family members also suffer from tinnitus), and common comorbidities (e.g., headaches, insomnia, hearing aids). Both questionnaires were used for the description of the sample.

Regarding the usage of app, users could set push notifications to on or off and were allowed to report their status at any time and as often as they wanted. Only time series datasets with at least 10 days of consecutive sampling were included in the analysis. More stringent cut-off criteria, such as 20 or 50 days of uninterrupted sampling, did not change the results obtained but decreased the sample size considerably (data not shown). If the same user had two sequences of observations that lasted at least 10 days, those two sequences were analyzed independently. Various strategies were explored when dealing with multiple entries from a given user in the same day (i.e., analyzing the mean or median values, the maximum or minimum values, or using the first or last observations). None of those methods to account for multiple observations on the same day changed the results (data not shown). The results reported in this article were obtained by selecting the first observation of each day. Missing values from a given sequence were imputed using the “aregImpute” function from the Hmisc package with default settings in R, after visually determining that missing data were likely missing at random.

### Statistical Analysis

Different statistical techniques were used to describe the relation between the variables collected with EMA, both at the group and individual levels. Autocorrelations and cross-correlations were used to obtain statistical associations at the group level, whereas linear regressions with elastic net regularization and unified structural equation modeling were used to obtain individualized models. These methods are described in the following.

### Auto-and Cross-Correlation

Autocorrelation can be described as the correlation of a variable with itself at different time lags. For this study, the time lag consisted of consecutive calendar days (see above). Mathematically, the autocorrelation, r_k_, can be expressed as:

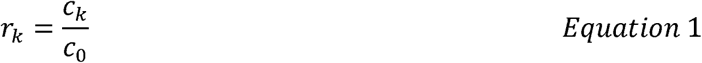

where C_0_ represents the autocovariance of a variable at lag 0, and C_k_ represents the autocovariance for lag k, which can be mathematically described as:

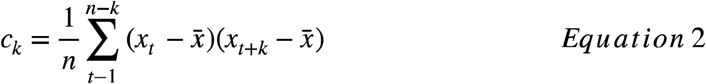

where *k* represents a lag (each lag representing one day), *t*, represents the *t*^*th*^ variable, and 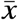 represents the mean of variable *x*. Similarly, cross-correlation can be described as the correlation between two variables at different lags and can be mathematically expressed as:

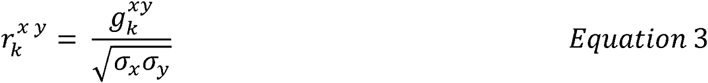

where sigma represents the standard deviation of variables *x* and *y*, and g represents the cross-correlation function, which can be represented as:

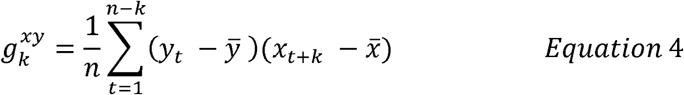

where *n* represents the sample size, *k* represents the lag, *t* represents the *t*^*th*^ variable, and both 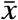 and 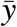 represents the mean of variables *x* and *y*. Auto-and cross-correlation were used to investigate whether the variables presented in Table 2 were associated with each other at the group level.

**Table 2.**
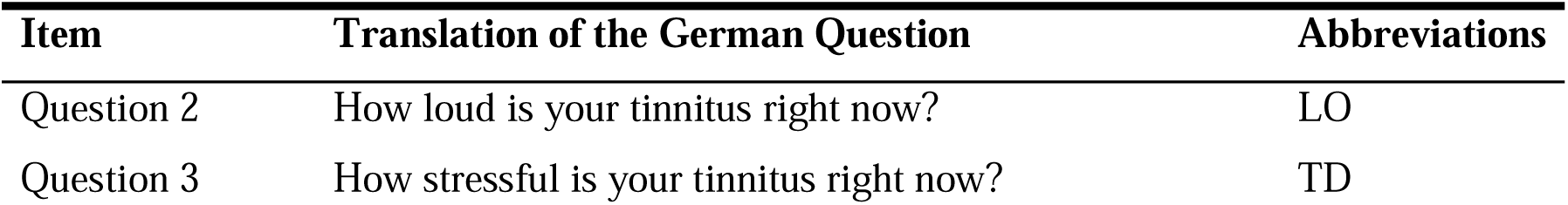
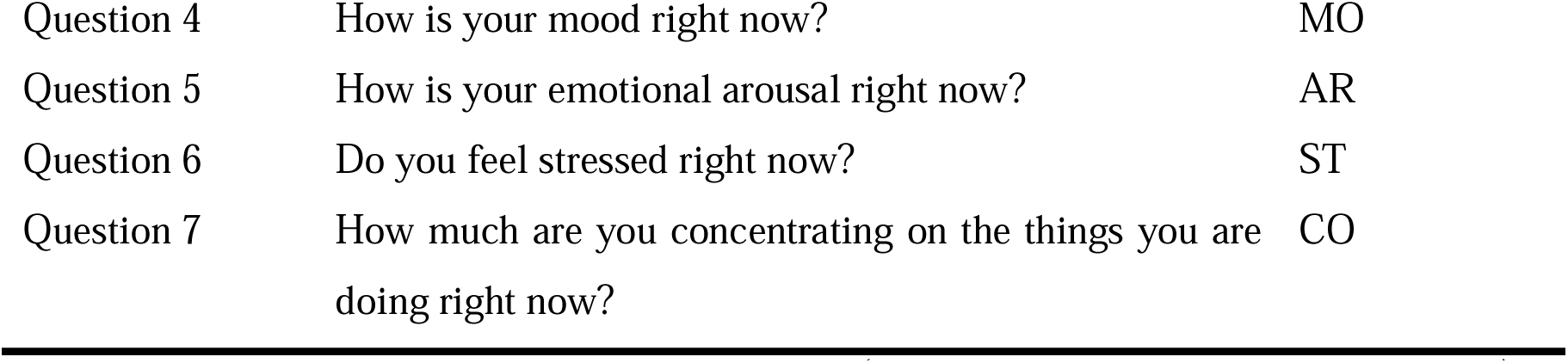
TYT questions included in the study. Question 1 (“Do you perceive your tinnitus right now?”) and question 8 (“Do you feel irritable right now?”) were excluded as their answers were dichotomous.

### Elastic Net Regularization

Elastic net regularization is an increasingly popular method intended to account for datasets with large numbers of predictors, especially when those may be correlated, which, in turn, may lead to overfitting of statistical models. The method combines two penalizing terms, *L1* and *L2*, and can be mathematically described as follows:

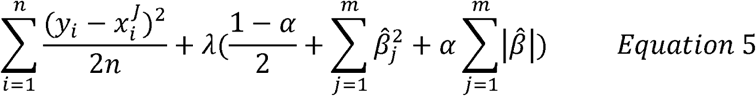

Where *n* is the sample size, *i* represents the *i*^*th*^ observation, j represents the *j*^*th*^ predictor, 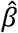 represents the estimated coefficients, lambda is the penalizing coefficient, and *α* is a tuning parameter: if set to 0, a ridge term is obtained, if set to 1, a lasso term is obtained. The first term represents the commonly used ordinal least squares (OLS) regression, the second represents an *L1* penalization (also known as ridge regression) and the third term represents an *L2* penalization (also known as lasso). If the lambda is estimated to 0, the output is a regular OLS regression. If not, the lambda must assume a positive value, in which the estimates from the regression are constrained. If a ridge penalization is used, the coefficients of regression are shrunk to values different than 0. Conversely, if a lasso regularization is used, the coefficients may be set to zero, which functions as a feature selection method. This powerful feature of the elastic net, to automatically select variables by setting some of them to 0, was used to build individualized models for each sequence of observations to predict both loudness (LO) and tinnitus distress (TD).

All variables in the regression were modeled as being linearly related to the outcome measures. A 10-fold cross-validation was computed to estimate lambda with the default settings using the cv.glmnet function. The final model was selected based on the lambda one standard error from the minimum for parsimonious results^22^.

### Unified Structural Equation Modelling

Unified Structural Equation Modeling (uSEM) combines structural equation modeling and vector autoregression and can be used to extract autoregressive and cross-lagged effects from time series. As a result, uSEM has been used widely in psychological and medical science to estimate contemporaneous and lagged effects from time series data (e.g., brain activity from functional magnetic resonance imaging and behavioral/emotional fluctuations recorded by EMA), both at the individual and group levels. The validity and reliability of idiographic methods from intensive longitudinal data sampling have been previously discussed^16^ and explored both with simulated and empirical data^23^.

uSEM estimates contemporaneous and lagged relations (of order Q) with the following formula:

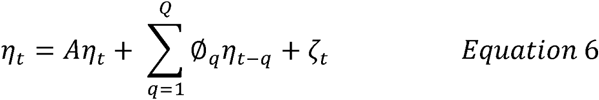

Where *η*_*t*_ the time series of length *t, A*, and Ø_q_ contain the matrix of dimensions (*p, p*) of the contemporaneous and lagged (at lag *q*) relations. S-GIMME was used to implement uSEM in our analysis, which is available as a package in R^24^.

All analyses were performed in R (version 4.0.1, R Core Team, 2018). Auto-and cross-correlations were conducted with an internal script adapted by JB from the functions ‘ACF’ and ‘CCF’ available in R. The adapted functions calculate weighted average auto-and cross correlations and therefore were applied to multiple sequences of observations.

## Results

Table 1 summarizes the demographics of the samples used for both analyses. Of the original dataset, 57% of the data were excluded from the analysis as the data were not obtained from that sequence for at least 10 uninterrupted days (see supplementary Figure 2). Thus, the sample of study 1 consisted of 488 unique sequences from 278 users). Following the guidelines of the authors of the s-GIMME package, only sequences with at least 60 days of uninterrupted usage were included in the second analysis^23^. Thus, the sample consisted of 32 sequences from 32 unique users. Table 2 shows how the EMA questions were formulated (translated into English from German), with their abbreviations which are used from here on. Two questions, namely questions 1 and 8, were excluded from the analysis as they were dichotomous and continuous variables were necessary for computing auto-and cross-correlations.

Figure 1 illustrates how uniquely tinnitus is experienced by four arbitrarily selected TYT users. The left column of the figure shows the mean values and their dispersion with kernel plots; the middle column shows the fluctuation of those variables through time with time series plots; the right column depicts the contemporaneous relation between variables with correlational heat maps. Overall, these four examples highlight how symptoms may be burdensome, how they fluctuate over time, and how they interact with each other.

**Figure 1.**
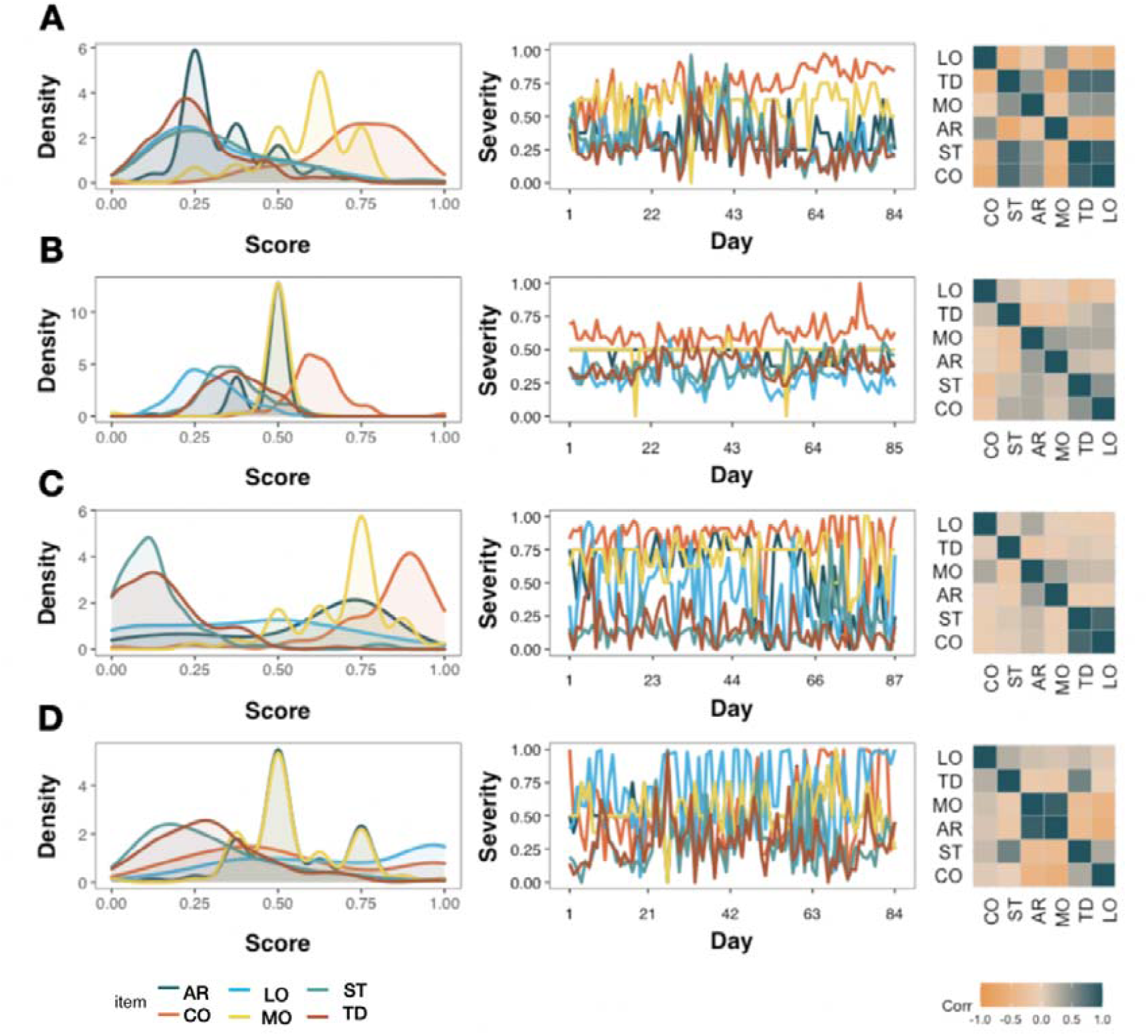
Variability between four TYT users, each represented in one row (A-D) who completed around 85 days of continuous EMA. Density plots (left), time series (middle) and correlation heat maps (right) highlight how arousal (AR), concentration (CO), loudness (LO), mood (MO), stress (ST) and TD (tinnitus distress) interact with each other.

### Auto and / or cross-correlations between tinnitus loudness, distress, and variables related to mood

First, we investigated whether the six variables were autocorrelated (see Figure 2). None of the lagged variables was outside the 95% confidence interval (red dashed lines), suggesting that there was no autocorrelation. Autocorrelations in lag 0 were always 1, since the nominators and denominators were identical in those cases (see Equation 1). Next, we investigated whether there was cross-correlation between the variables (see Figure 3). Like the previous results, no correlation at lags > 0 was observed. However, corroborating previous findings, we observed contemporaneous correlations (i.e., at lag 0) between LO & TD, TD & MO, LO & ST, TD & ST, MO & AR, MO & ST and AR & ST.

**Figure 2.**
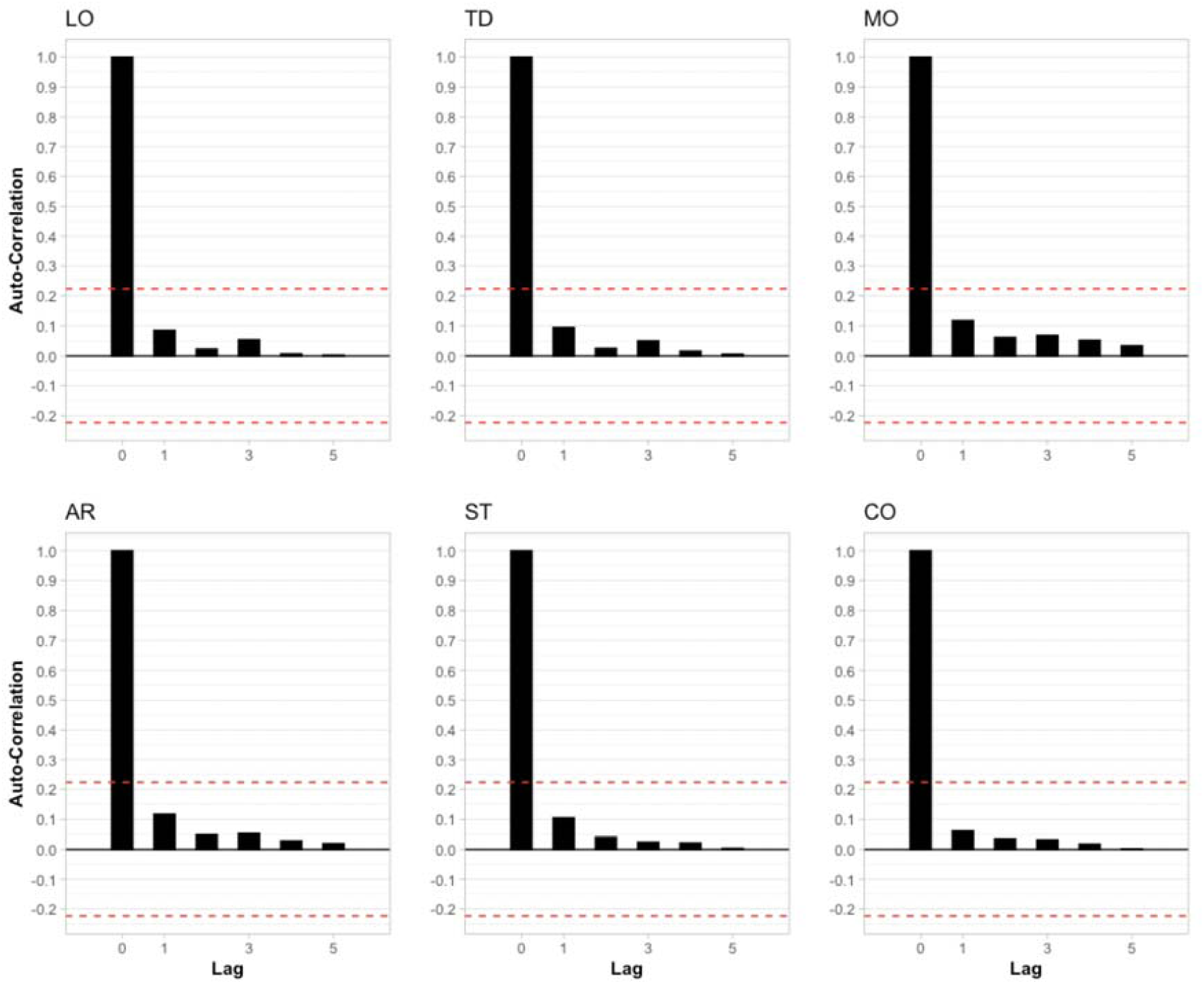
Autocorrelation of the six variables included in the analysis. Each lag represents a different day of usage. Red dashed lines represent the 95% confidence interval. LO: Loudness; TD: Tinnitus Distress; MO: Mood; AR: Arousal; ST: Stress; CO: Concentration

**Figure 3.**
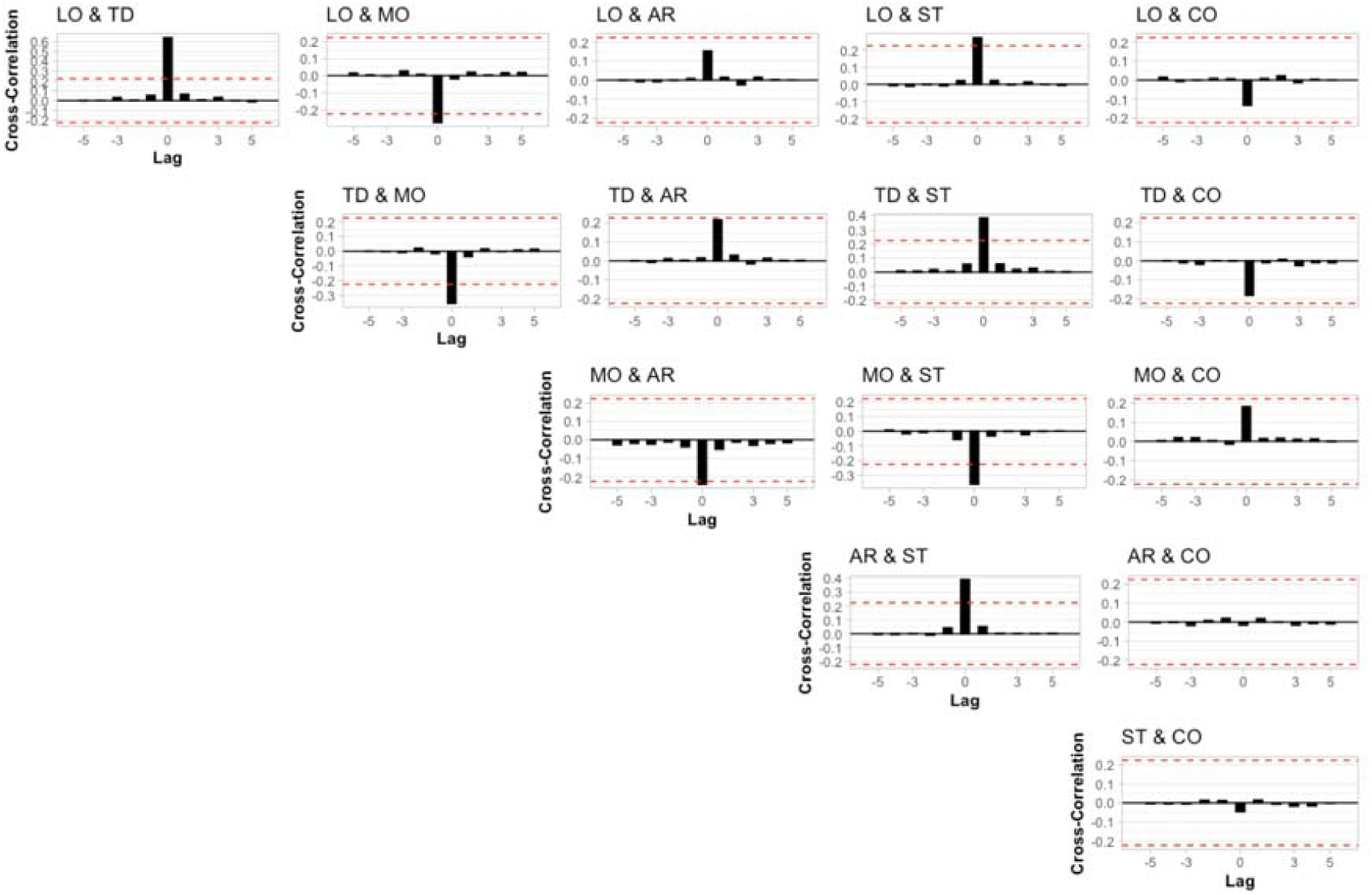
Cross-correlation of the potential combinations of the six variables included in the analysis. Each lag represents a different day of usage. Red dashed lines represent the 95% confidence interval. LO: Loudness; TD: Tinnitus Distress; MO: Mood; AR: Arousal; ST: Stress; CO: Concentration

### Individualized Models with Elastic Net Regularization

Next, we investigated whether elastic net regressions could be used for individualized inference about LO (see Figures 4A-C) and TD (see Figures 4D-F). For this analysis, contemporaneous variables and lagged variables from the previous days (acronyms ending with “1” in Figures 4A, 4C, 4D and 3F) were used as independent variables in regression setups. For 27% and 31% of the sample, no predictors of LO and TD were found (see Figures 4A and 4D, respectively). For the remaining sample, the R^2^ for each time series varied considerably (see Figures 4B and 4E). Figures 4C and 4F summarize these findings with box plots. Although certain variables were almost only positively associated with the outcome measure (e.g., LO, TD and ST), other variables presented both positive and negative valence throughout the sample (e.g., CO and LO).

**Figure 4.**
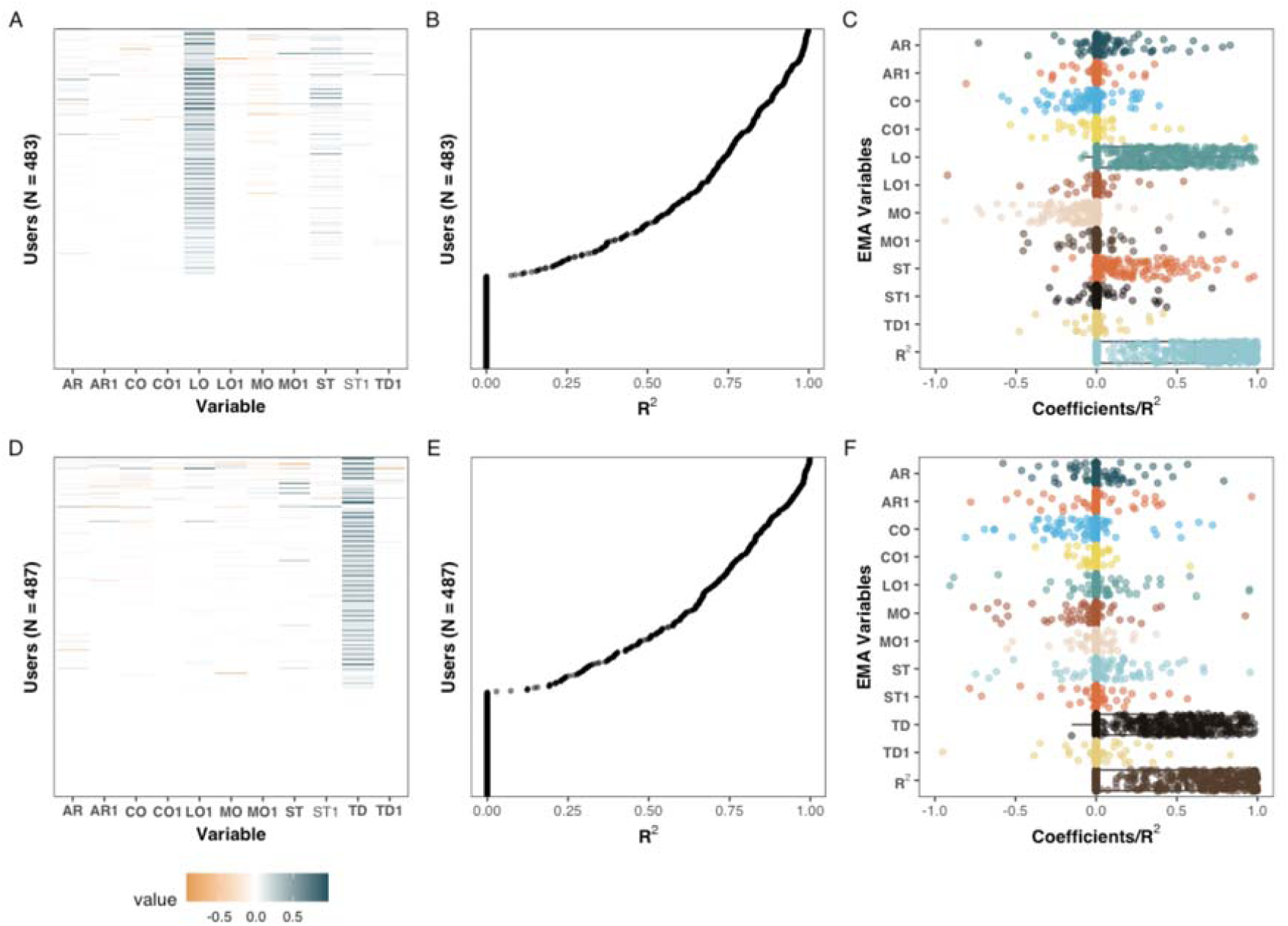
Coefficients of penalized regression models with TD (A-C) and LO (D-F) as dependent variables. Sequences in which the dependent variable had no variability were excluded from this analysis. The coefficients and R^2^ of the individualized models on the Y-axis of Figures 3A and 3B, and of Figures 3D and 3E are aligned. Left column: standardized coefficients uniquely estimated for each sequence. Middle column: the amount of variance explained, measured with R^2^, for each unique sequence. Right column: The adjusted coefficients of A and D, and R^2^, of B and E are presented with box and dot plots.

Based on the model coefficients, positive associations were observed between LO and TD among 314 users (65%) at the contemporaneous level and 17 (3.52%) users at the lagged level. ST was positively associated with TD for 133 users (27.5%) and MO was negatively associated with TD in 105 (21.7) cases. The associations between TD and the remaining variables, positive or negative, were present only among a fraction of users (0.6% - 11.4%). Apart from the association between LO and TD, few associations with LO (0.2% - 11%) were observed. ST was positively associated with LO for 53 (11%) of the users, and MO was negatively associated with 50 users (10.4%).

### Idiographic Modeling with S-GIMME

Only time series with at least 60 sequential observations were included in this analysis, yielding a sample size of 32 unique users. The models converged in all cases and a good fit was observed: (average: *X*^2^ (44, *N* = 32) = 65.56, *p* = 0.12, CFI = 0.98, RMSEA = 0.06, NNFI = 0.96, SRMR = 0.06). Individual model fits are available as supplemental material.

Figure 5 shows the paths for the four individuals from Figure 1. The variable “Day”, that is, the position in the time series was encoded as an exogenous variable, meaning that the variable could predict any other variables, but not the other way around. Both contemporaneous (solid) and lagged (dashed) paths were obtained in all cases, including the four highlighted cases in Figure 5. Although some paths were shared across subjects, e.g., the effect of TD on LO, other dynamics were idiographic (e.g., whereas ST had a positive contemporaneous effect on TD for user shown in Figure 5A, the relationship was inverted for the user shown in Figure 5D, and no relationship was seen for the other two users).

**Figure 5.**
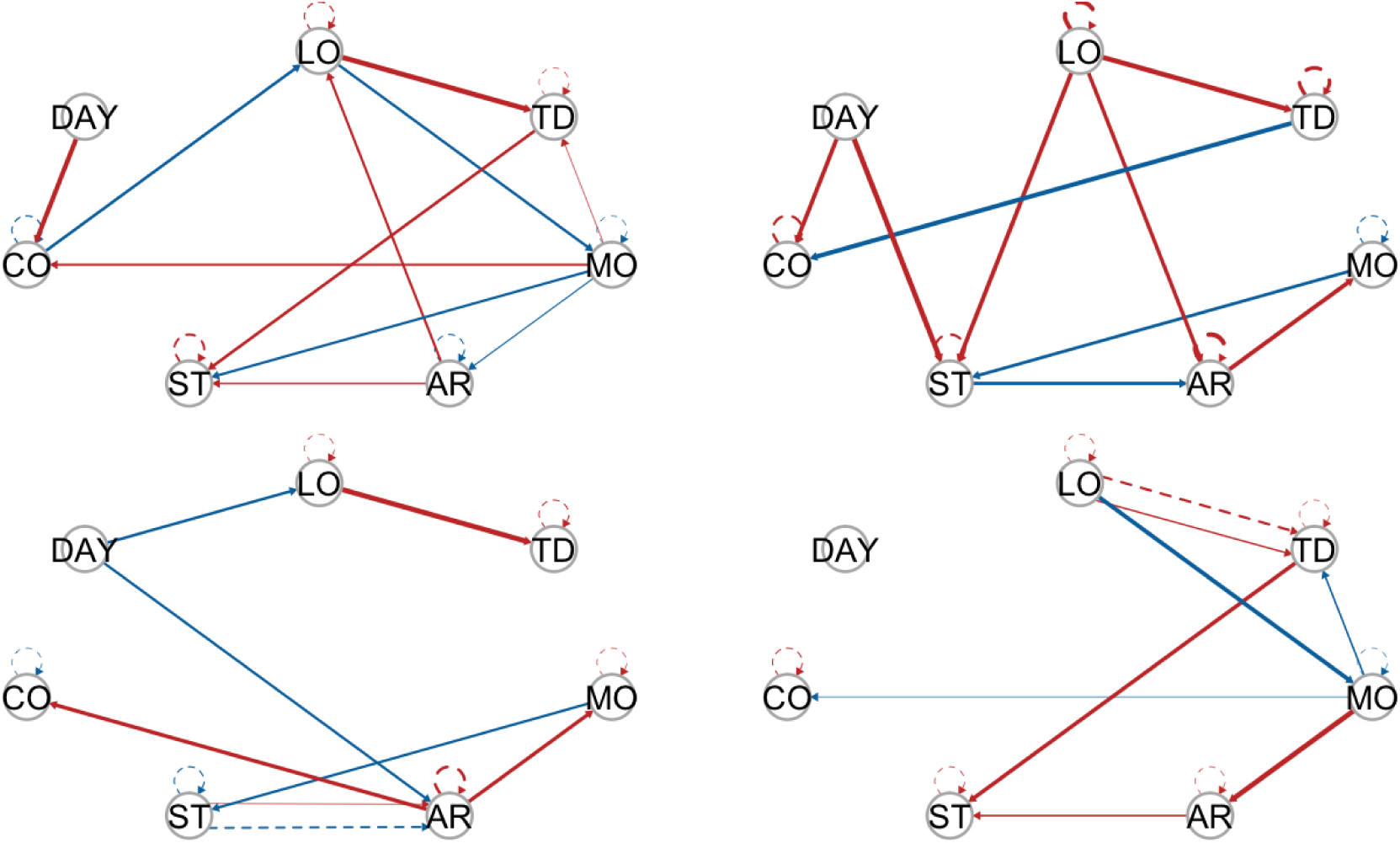
Model estimated by S-GIMME by the same four individuals from Figure 1. Solid lines represent a contemporaneous effect (lag = 0), and dashed lines represent lagged effects (lag = 1). The circles represent the variables included in the study. Arrows indicate the direction of the relationship. Red arrows indicate positive regression paths and blue lines indicate negative regression paths.

S-GIMME identified two subgroups based on the dynamic similarities between variables collected through EMA. Group 1 consisted of 19 individuals, while group 2 consisted of 13 individuals. No differences were observed between the subgroups in terms of mean EMA scores, sociodemographics, or tinnitus characteristics (see Table 3). The paths obtained from S-GIMME for each subgroup are shown in Figure 6. Interestingly, most of the lagged relations were observed in subgroup 1 (ie., dashed lines), whereas subgroup 2 consisted mainly of contemporaneous (ie., solid lines), suggesting that S-GIMME could distinguish users whose tinnitus is mostly modulated from contemporaneous effects from users whose tinnitus is modulated by dynamics from the previous day. The green paths shown in Figure 6 highlight effects specific to all members of a subgroup. Of interest, most of the effects specific for subgroup 2 originated from either TD or LO. These findings suggest that S-GIMME could not only distinguish users with contemporaneous or lagged dynamics, but also users whose TD and LO were associated with another, and with AR, ST and MO (see Figure6, right).

**Table 3.**
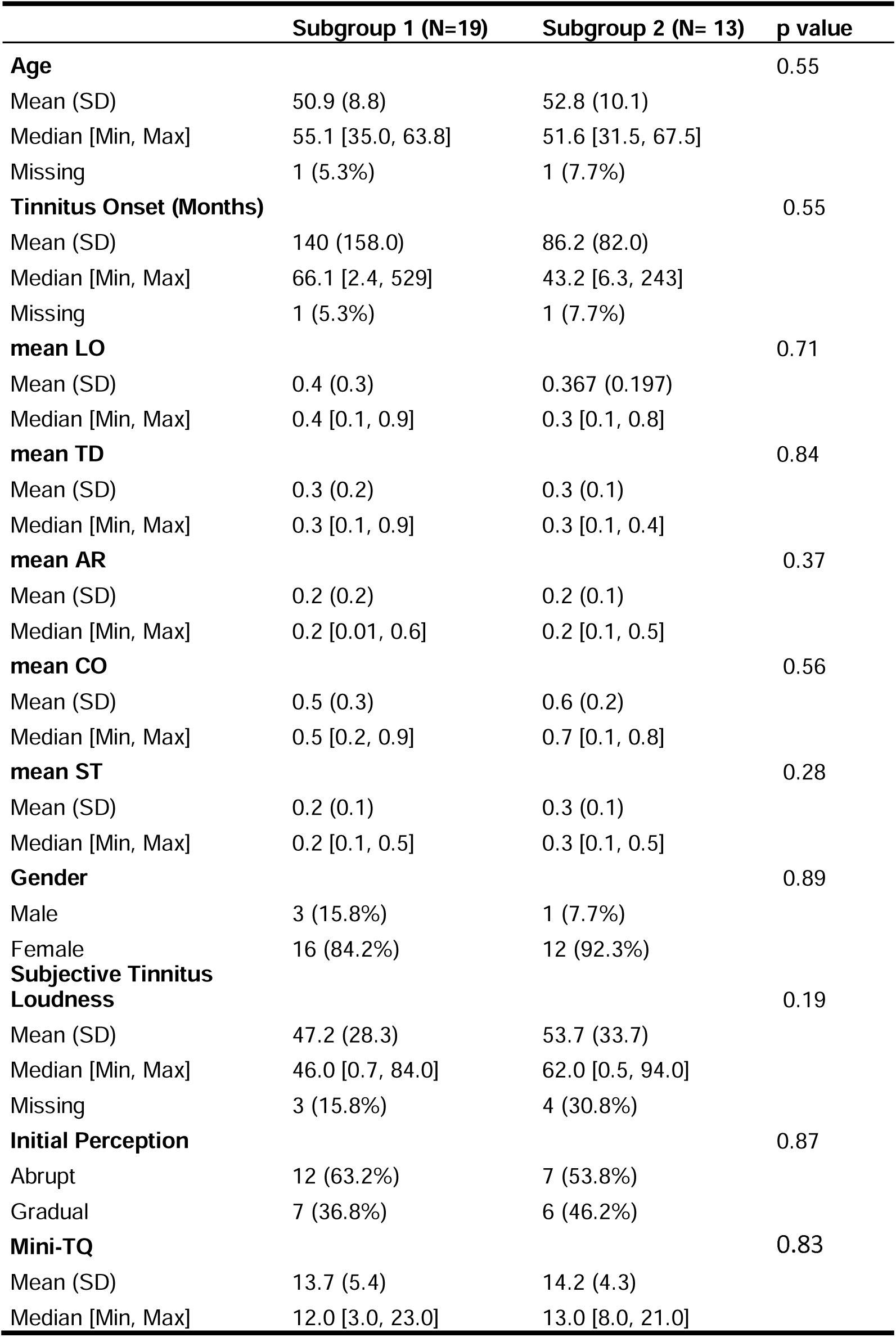
Description of the 32 users included in the analysis with S-GIMME. P values were obtained from the Kruskal-Wallis test if the response variable was continuous or from Chi-square tests if the response variable was categorical. No corrections were made for multiple comparisons.

**Figure 6.**
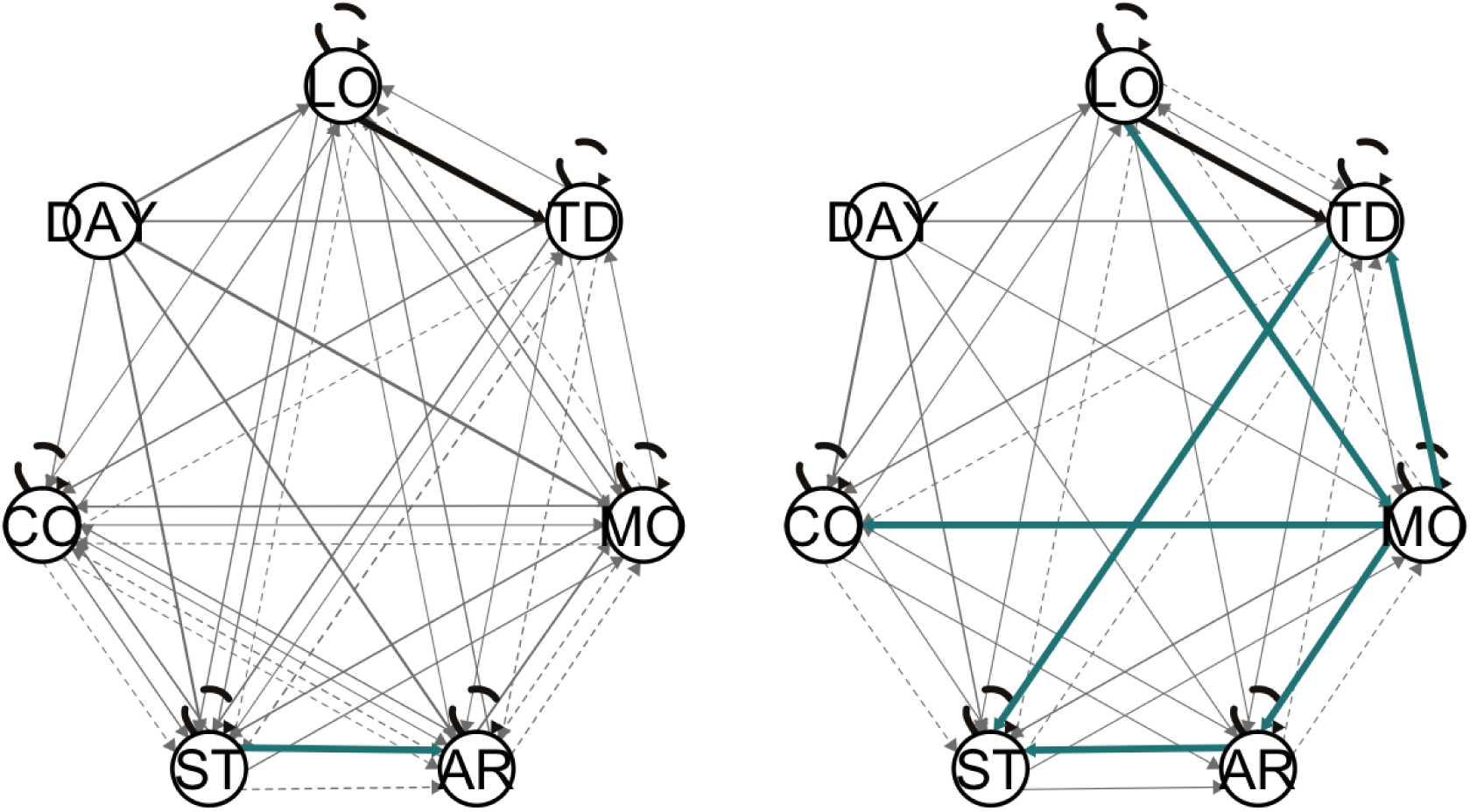
Two subgroups were identified by S-GIMME (left: N = 19, right: N = 13). The black lines represent the paths present at the group level, the green lines represent the paths present at the subgroup level, and the gray lines represent the paths at the individual level. Solid lines represent contemporaneous effects, and dashed lines represent lagged effects. Arrow heads indicate the direction of the relationship.

## Discussion

In this study, we identified individuals whose tinnitus loudness and distress are affected by behavioral and emotional processes of the same and of the previous day. We started by showing that at the group level, no evidence of autocorrelation or cross-correlation could be observed between the six EMA variables. However, modeling data at the individual level revealed that LO and TD were auto-and cross-lagged from one day to another for several individuals (see Figures 3, 4 and 5). Lastly, we used S-GIMME to explore the unique interplay of these variables, effectively modeling how heterogeneously tinnitus manifests itself over time.

Our results provide a template for how to model LO and TD at the individual level (see Figures 3, 4, and 5). The interaction between these variables constitutes a complex mosaic of how tinnitus is experienced; Although the uniqueness of experiencing tinnitus has been widely acknowledged^7,15,25^, empirical studies that demonstrate this complex relationship were lacking. Furthermore, using S-GIMME, we were able to incorporate the effect of time into the models, a critical component of experiencing tinnitus^26^ often neglected in empirical studies due to the challenges of conducting multiple samplings in the traditional ambulatory setting.

Corroborating previous findings, we showed a positive association between ST, LO, and TD^17,27^. Interestingly, we also observed that AR, MO and CO had ambivalent associations with both loudness and distress (see Figures 3C and 3F). Such associations could explain why tinnitus is uniquely experienced, and future research could further investigate factors associated with TD and LO, such as behavior, emotional and cognitive dynamics samples from EMA. Tinnitus is known to have potential negative consequences on cognition^28–30^, but this is the first time a positive association between CO and TD / LO has been shown. Future studies should further investigate this seemingly paradoxical relationship, especially considering that concentration problems are one of the core domains that tinnitus patients would like to have as outcome measures from clinical interventions^31,32^. For example, this positive association could be related to compensatory mechanisms.

Other core domains include the ability to ignore the phantom perception, its intrusiveness, the sense of control over one’s body, quality of sleep, and negative thoughts and beliefs. The first three domains could be investigated by adapting questions from the Tinnitus Functional Index, a widely used, validated questionnaire that captures different dimensions of tinnitus distress^33^. Negative thoughts could be investigated by adapting the positive and negative affect schedule^34^, similar to other studies in the field of psychopathology using EMA^16,35^. Sleep problems remain one of the main complaints of clinical tinnitus patients^7,36–39^, but no study to date investigated its effect using an EMA design, despite the evidence of its relevance on other chronic, disabling conditions^40^.

Our work suggests that only a subset of patients experience lagged effects between the variables sampled through EMA (see Figures 4, 5, and 6). This finding could be leveraged to deliver personalized interventions through mobile apps, especially among users where TD and LO and auto or cross-correlated. For example, the results suggest that patients with tinnitus may benefit from just-in-time adaptive intervention (JITAI)^41,42^. JITAI uses mobile sensing to deliver customized interventions based on unique fluctuations recorded by the EMA. Such a system has been used in several fields, including physical health, addiction, and mental care research, but not in tinnitus^41^. For example, customized interventions could be delivered to patients whose LO and/or TD are auto-or cross-correlated over days: Once the algorithm anticipates a potential spike in LO or ST on the next day, an intervention such as psychoeducation tips for coping with tinnitus through push notifications^43^, sound therapy^44–46^, online delivered cognitive behavior techniques^13,47,48^, or meditation techniques^49^ could be activated. Future studies could evaluate whether these results are replicated with different sampling frequencies. For example, an EMA every 8 hours has been shown to be well tolerated in clinical settings^50^ and would considerably reduce the duration of the study, and potentially increase the adherence to using the APP^51^.

Methodological limitations should be considered when interpreting these results. For example, missing values continue to constitute a significant challenge for researchers, including those using EMA. A recent study investigated the causes of discontinuation of TYT use^51^, but no clear predictors of adherence to app use were found. Furthermore, results suspected of bias cannot be easily discarded, as only a fraction of users used the app for more than 10 consecutive days (supplemental material). In this study, we implemented a popular and robust method for data imputation; however, empirical evidence that this is the optimal method for imputing data from EMA is not available. Finally, the possibility of low stability from the coefficients obtained from elastic net should also be considered, especially when covariates are correlated^52^. Future studies could attenuate the effects of measurement error by composing latent factors with S-GIMME.

In summary, we show that tinnitus loudness and tinnitus distress are affected by both contemporaneous and lagged behavioral and emotional processes measured through EMA. Additionally, we showed that S-GIMME can distinguish users whose tinnitus distress and loudness are modulated by contemporaneous effects from those where those dynamics are modulated by both contemporaneous and lagged effects. Distinguishing these two subgroups could be therapeutical value, especially when aligned with just-in-time adaptive interventions to mitigate or prevent future peaks of tinnitus distress or increased loudness.

## Data Availability

All data produced in the present study are available upon reasonable request to the authors

## References

1. Baguley, D., McFerran, D. & Hall, D. Tinnitus. Lancet 382, 1600–1607 (2013).

2. Langguth, B., Kreuzer, P. M., Kleinjung, T. & Ridder, D. D. Tinnitus: causes and clinical management. Lancet Neurology 12, 920–930 (2013).

3. Shore, S. E., Roberts, L. E. & Langguth, B. Maladaptive plasticity in tinnitus — triggers, mechanisms and treatment. Nat Rev Neurol 12, 150–160 (2016).

4. Biswas, R. & Hall, D. A. The Behavioral Neuroscience of Tinnitus. Curr Top Behav Neurosci 51, 3–28 (2020).

5. Cima, R. F. F. Bothersome tinnitus. Hno 66, 369–374 (2018).

6. Maes, I. H. L., Cima, R. F. F., Vlaeyen, J. W., Anteunis, L. J. C. & Joore, M. A. Tinnitus. Ear Hearing 34, 508–514 (2013).

7. Tunkel, D. E. et al. Clinical Practice Guideline. Otolaryngology Head Neck Surg 151, S1– S40 (2014).

8. Simões, J. P., Neff, P. K. A., Langguth, B., Schlee, W. & Schecklmann, M. The progression of chronic tinnitus over the years. Sci Rep-uk 11, 4162 (2021).

9. Kleinjung, T. & Langguth, B. Avenue for Future Tinnitus Treatments. Otolaryng Clin N Am 53, 667–683 (2020).

10. Simoes, J. et al. Toward Personalized Tinnitus Treatment: An Exploratory Study Based on Internet Crowdsensing. Frontiers Public Heal 7, 157 (2019).

11. Genitsaridi, E., Hoare, D. J., Kypraios, T. & Hall, D. A. A Review and a Framework of Variables for Defining and Characterizing Tinnitus Subphenotypes. Brain Sci 10, 938 (2020).

12. Maas, I. L. et al. Genetic susceptibility to bilateral tinnitus in a Swedish twin cohort. Genet Med 19, 1007–1012 (2017).

13. Kleinstäuber, M., Weise, C., Andersson, G. & Probst, T. Personality traits predict and moderate the outcome of Internet-based cognitive behavioural therapy for chronic tinnitus. Int J Audiol 57, 1–7 (2018).

14. Hafner, A. et al. Impact of personality on acoustic tinnitus suppression and emotional reaction to stimuli sounds. Prog Brain Res 260, 187–203 (2020).

15. Cederroth, C. R. et al. Editorial: Towards an Understanding of Tinnitus Heterogeneity. Front Aging Neurosci 11, 53 (2019).

16. Wright, A. G. C. & Woods, W. C. Personalized Models of Psychopathology. Annu Rev Clin Psycho 16, 1–26 (2020).

17. Probst, T., Pryss, R., Langguth, B. & Schlee, W. Emotional states as mediators between tinnitus loudness and tinnitus distress in daily life: Results from the “TrackYourTinnitus” application. Sci Rep-uk 6, 20382 (2016).

18. Pryss, R. et al. Prospective crowdsensing versus retrospective ratings of tinnitus variability and tinnitus–stress associations based on the TrackYourTinnitus mobile platform. Int J Data Sci Anal 8, 327–338 (2019).

19. Schlee, W. et al. Measuring the Moment-to-Moment Variability of Tinnitus: The TrackYourTinnitus Smart Phone App. Front Aging Neurosci 8, 294 (2016).

20. Hiller, W. & Goebel, G. Rapid assessment of tinnitus-related psychological distress using the Mini-TQ. International Journal of Audiology 600–604 (4AD).

21. Langguth, B. et al. Consensus for tinnitus patient assessment and treatment outcome measurement: Tinnitus Research Initiative meeting, Regensburg, July 2006. Prog Brain Res 166, 525–536 (2007).

22. Zou, H. & Hastie, T. Regularization and variable selection via the elastic net. J Royal Statistical Soc Ser B Statistical Methodol 67, 301–320 (2005).

23. Lane, S. T., Gates, K. M., Pike, H. K., Beltz, A. M. & Wright, A. G. C. Uncovering General, Shared, and Unique Temporal Patterns in Ambulatory Assessment Data. Psychol Methods 24, 54–69 (2019).

24. Lane, S. T. & Gates, K. M. Automated Selection of Robust Individual-Level Structural Equation Models for Time Series Data. Struct Equ Model Multidiscip J 24, 1–15 (2017).

25. Elgoyhen, A. B., Langguth, B., Ridder, D. D. & Vanneste, S. Tinnitus: perspectives from human neuroimaging. Nat Rev Neurosci 16, 632–642 (2015).

26. Probst, T. et al. Does Tinnitus Depend on Time-of-Day? An Ecological Momentary Assessment Study with the “TrackYourTinnitus” Application. Front Aging Neurosci 9, 253 (2017).

27. Scott, B., Lindberg, P., Melin, L. & Lyttkens, L. Predictors of tinnitus discomfort, adaptation and subjective loudness. Brit J Audiol 24, 51–62 (2009).

28. Andersson, G. & McKenna, L. The role of cognition in tinnitus. Acta Oto-laryngol 126, 39–43 (2009).

29. Mohamad, N., Hoare, D. J. & Hall, D. A. The consequences of tinnitus and tinnitus severity on cognition: A review of the behavioural evidence. Hearing Res 332, 199–209 (2016).

30. Neff, P. et al. The impact of tinnitus distress on cognition. Sci Rep-uk 11, 2243 (2021).

31. Hall, D. A. et al. The COMiT’ID Study: Developing Core Outcome Domains Sets for Clinical Trials of Sound-, Psychology-, and Pharmacology-Based Interventions for Chronic Subjective Tinnitus in Adults. Trends Hear 22, 2331216518814384 (2018).

32. Hall, D. A. et al. One Size Does Not Fit All: Developing Common Standards for Outcomes in Early-Phase Clinical Trials of Sound-, Psychology-, and Pharmacology-Based Interventions for Chronic Subjective Tinnitus in Adults. Trends Hear 23, 2331216518824827 (2019).

33. Meikle, M. B. et al. The Tinnitus Functional Index. Ear Hearing 33, 153–176 (2012).

34. Watson, D., Clark, L. A. & Tellegen, A. Development and Validation of Brief Measures of Positive and Negative Affect: The PANAS Scales. J Pers Soc Psychol 54, 1063–1070 (1988).

35. Heller, A. S., Stamatis, C. A., Puccetti, N. A. & Timpano, K. R. The distribution of daily affect distinguishes internalizing and externalizing spectra and subfactors. J Abnorm Psychol 130, 319–332 (2021).

36. Asnis, G. M. et al. An Examination of the Relationship Between Insomnia and Tinnitus: A Review and Recommendations. Clin Medicine Insights Psychiatry 9, 1179557318781078 (2018).

37. Richter, K. et al. Insomnia Associated with Tinnitus and Gender Differences. Int J Environ Res Pu 18, 3209 (2021).

38. Inagaki, Y. et al. Personality and sleep evaluation of patients with tinnitus in Japan. (2020) doi:10.21203/rs.3.rs-30270/v1.

39. Lu, T. et al. Positive Correlation between Tinnitus Severity and Poor Sleep Quality Prior to Tinnitus Onset: a Retrospective Study. Psychiat Quart 91, 379–388 (2020).

40. Short, N. A., Allan, N. P. & Schmidt, N. B. Sleep disturbance as a predictor of affective functioning and symptom severity among individuals with PTSD: An ecological momentary assessment study. Behav Res Ther 97, 146–153 (2017).

41. Wang, L. & Miller, L. C. Just-in-the-Moment Adaptive Interventions (JITAI): A Meta-Analytical Review. Health Commun 35, 1–14 (2019).

42. Nahum-Shani, I. et al. Just-in-Time Adaptive Interventions (JITAIs) in Mobile Health: Key Components and Design Principles for Ongoing Health Behavior Support. Ann Behav Med 52, 1–17 (2016).

43. Unnikrishnan, V. et al. The Effect of Non-Personalised Tips on the Continued Use of Self-Monitoring mHealth Applications. Brain Sci 10, 924 (2020).

44. Kutyba, J. et al. Effectiveness of tinnitus therapy using a mobile application. Eur Arch Oto-rhino-l 1–11 (2021) doi:10.1007/s00405-021-06767-9.

45. Tyler, R. S. et al. Tinnitus Suppression in Cochlear Implant Patients Using a Sound Therapy App. Am J Audiol 27, 1–8 (2018).

46. Kutyba, J., Jedrzejczak, W. W., Raj-Koziak, D., Gos, E. & Skarzynski, P. H. TINNITUS SOUND THERAPY WITH A MOBILE APPLICATION: CASE STUDY. J Hear Sci 9, 51– 56 (2019).

47. Andersson, G., Titov, N., Dear, B. F., Rozental, A. & Carlbring, P. Internet□delivered psychological treatments: from innovation to implementation. World Psychiatry 18, 20–28 (2019).

48. Weise, C., Kleinstäuber, M. & Andersson, G. Internet-Delivered Cognitive-Behavior Therapy for Tinnitus. Psychosom Med 78, 501–510 (2016).

49. Fitzgerald, B. P., Stocking, C., Ralli, M. & Sheppard, A. At-home meditation for tinnitus management. Hear Balance Commun 19, 1–9 (2021).

50. Shiffman, S., Stone, A. A. & Hufford, M. R. Ecological Momentary Assessment. Annu Rev Clin Psycho 4, 1–32 (2008).

51. Schleicher, M. et al. Understanding adherence to the recording of ecological momentary assessments in the example of tinnitus monitoring. Sci Rep-uk 10, 22459 (2020).

52. Shen, Y., Han, B. & Braverman, E. Stability of the elastic net estimator. J Complexity 32, 20–39 (2016).

