## Supplemental Figure and Table 1 for "Tinnitus Subtyping with Subgrouping Within Group Iterative Multiple Model Estimation: An Ecological Momentary Assessment Study"

Supplemental material

Affiliations:

1) Department of Psychiatry and Psychotherapy, University of Regensburg, Regensburg, Germany
2) Department of Mathematics, University of Bergen, Bergen, Norway
3) Institute of Bioengineering, Center for Neuroprosthetics, École Polytechnique Fédérale de Lausanne, Switzerland
4) Department of Radiology and Medical Informatics, University of Geneva, Switzerland
5) Center for Cognitive Neuroscience (CCNS) and Department of Psychology, University of Salzburg, Salzburg, Austria
6) Institute of Clinical Epidemiology and Biometry, University of Würzburg, Würzburg, Germany
7) Department of Otolaryngology, Regensburg University Hospital, Regensburg, Germany

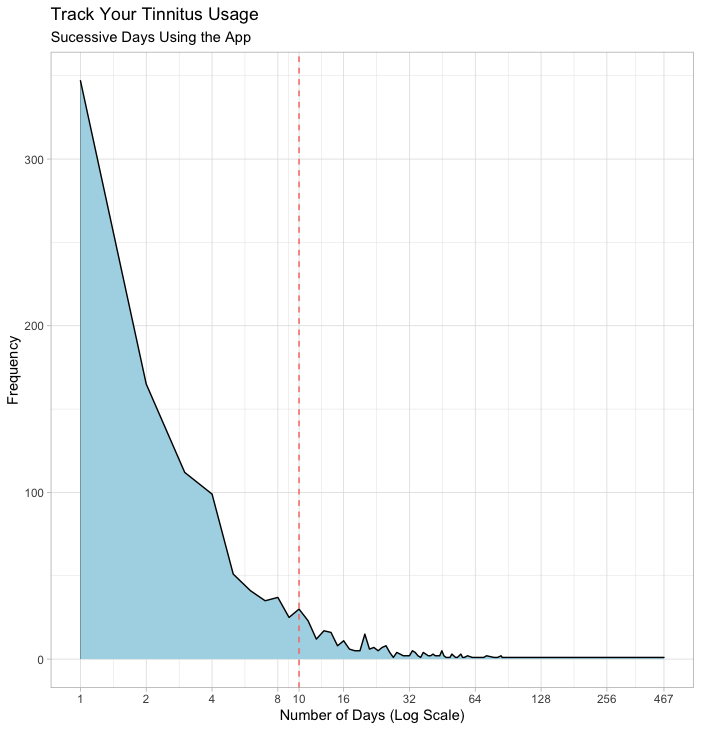

Sup. Figure 1 Overview of data. The x-axis shows the length of consecutive days in a logarithm scale ranging from one to 467 days, and the y-axis shows how frequent that sequence of consecutive days was observed. The red vertical dashed line shows the cut-off point of at least 10 days of consecutive observations that was used as inclusion criteria.

| User | chisq | df | npar | pvalue | rmsea | srmr | nnfi | cfi | bic | aic | logl |
| --- | --- | --- | --- | --- | --- | --- | --- | --- | --- | --- | --- |
| 1 | 63.96 | 45 | 59 | 0.03 | 0.05 | 0.06 | 0.97 | 0.98 | 5431.34 | 5244.96 | -2563.48 |
| 2 | 54.95 | 45 | 59 | 0.15 | 0.06 | 0.08 | 0.95 | 0.97 | 2040.61 | 1917.05 | -899.52 |
| 3 | 68.46 | 42 | 62 | 0.01 | 0.09 | 0.05 | 0.94 | 0.97 | 2480.38 | 2330.42 | -1103.21 |
| 4 | 55.06 | 41 | 63 | 0.07 | 0.05 | 0.05 | 0.93 | 0.96 | 5579.62 | 5388.3 | -2631.15 |
| 5 | 42.57 | 39 | 65 | 0.32 | 0.04 | 0.07 | 0.96 | 0.98 | 2386.66 | 2246.33 | -1058.17 |
| 6 | 68.88 | 45 | 59 | 0.01 | 0.06 | 0.07 | 0.96 | 0.98 | 3912.58 | 3744.77 | -1813.39 |
| 7 | 57.7 | 44 | 60 | 0.08 | 0.06 | 0.06 | 0.96 | 0.98 | 2518.83 | 2375.91 | -1127.96 |
| 8 | 72.41 | 45 | 59 | 0.01 | 0.07 | 0.06 | 0.96 | 0.97 | 4008.64 | 3839.46 | -1860.73 |
| 9 | 84.04 | 48 | 56 | 0 | 0.05 | 0.03 | 0.94 | 0.97 | 11988.77 | 11773.53 | -5830.77 |
| 10 | 47.22 | 42 | 62 | 0.27 | 0.04 | 0.07 | 0.97 | 0.98 | 3055.64 | 2904.93 | -1390.46 |
| 11 | 50.08 | 43 | 61 | 0.21 | 0.04 | 0.06 | 0.96 | 0.98 | 3033.89 | 2884.17 | -1381.09 |
| 12 | 56.69 | 40 | 64 | 0.04 | 0.05 | 0.05 | 0.96 | 0.98 | 5811.48 | 5610.79 | -2741.4 |
| 13 | 53.46 | 43 | 61 | 0.13 | 0.05 | 0.07 | 0.95 | 0.97 | 3018.87 | 2869.16 | -1373.58 |
| 14 | 53.33 | 46 | 58 | 0.21 | 0.05 | 0.07 | 0.95 | 0.97 | 2434.45 | 2306.58 | -1095.29 |
| 15 | 47.65 | 43 | 61 | 0.29 | 0.04 | 0.07 | 0.98 | 0.99 | 2010.34 | 1881.58 | -879.79 |
| 16 | 58.28 | 46 | 58 | 0.11 | 0.06 | 0.07 | 0.97 | 0.98 | 2215.59 | 2084.36 | -984.18 |
| 17 | 67.7 | 40 | 64 | 0 | 0.04 | 0.04 | 0.96 | 0.98 | 12272.59 | 12023.71 | -5947.85 |
| 18 | 66.34 | 42 | 62 | 0.01 | 0.08 | 0.06 | 0.95 | 0.97 | 2501.84 | 2348.24 | -1112.12 |
| 19 | 50.08 | 42 | 62 | 0.18 | 0.04 | 0.06 | 0.92 | 0.96 | 3919.21 | 3754.66 | -1815.33 |
| 20 | 53.33 | 44 | 60 | 0.16 | 0.06 | 0.08 | 0.96 | 0.98 | 2299.09 | 2166.81 | -1023.41 |
| 21 | 62.3 | 42 | 62 | 0.02 | 0.08 | 0.05 | 0.96 | 0.98 | 2209.39 | 2064.07 | -970.04 |
| 22 | 45.81 | 40 | 64 | 0.24 | 0.05 | 0.07 | 0.96 | 0.98 | 2169.26 | 2034.17 | -953.08 |
| 23 | 75.95 | 43 | 61 | 0 | 0.06 | 0.08 | 0.95 | 0.97 | 6906.86 | 6701.53 | -3289.77 |
| 24 | 48.92 | 43 | 61 | 0.25 | 0.04 | 0.06 | 0.97 | 0.98 | 2931.34 | 2783.79 | -1330.9 |
| 25 | 50.72 | 40 | 64 | 0.12 | 0.05 | 0.06 | 0.96 | 0.98 | 3016.32 | 2855.63 | -1363.81 |
| 26 | 55.39 | 45 | 59 | 0.14 | 0.06 | 0.08 | 0.95 | 0.97 | 2323.8 | 2191.99 | -1036.99 |
| 27 | 51.34 | 44 | 60 | 0.21 | 0.05 | 0.07 | 0.96 | 0.98 | 2190.08 | 2061.5 | -970.75 |
| 28 | 125.64 | 43 | 61 | 0 | 0.09 | 0.05 | 0.94 | 0.97 | 6447.8 | 6235.73 | -3056.87 |
| 29 | 62.99 | 44 | 60 | 0.03 | 0.07 | 0.05 | 0.97 | 0.98 | 2172.3 | 2030.13 | -955.06 |
| 30 | 190.92 | 45 | 59 | 0 | 0.18 | 0.04 | 0.93 | 0.96 | 393.07 | 242.39 | -62.2 |
| 31 | 56.17 | 43 | 61 | 0.09 | 0.05 | 0.05 | 0.96 | 0.98 | 4111.59 | 3941.04 | -1909.52 |
| 32 | 67.18 | 46 | 58 | 0.02 | 0.08 | 0.09 | 0.96 | 0.97 | 1794.71 | 1668.6 | -776.3 |

Sup. Table 1 Individual model fits from GIMME. All models converged normally. Chisq = Chi-squared, df = degrees of freedom, npar = number of parameters, rmsea = Root Mean Square Error of Approximation, srmr = Standardized Root Mean Square Residual, aic = Akaike information criterion, bic = Bayesian information criterion, logl = loglikelihood.
